# Data-driven discovery of changes in clinical code usage over time: a case-study on changes in cardiovascular disease recording in two English electronic health records databases (2001-2015)

**DOI:** 10.1101/19006098

**Authors:** Patrick Rockenschaub, Vincent Nguyen, Rob W. Aldridge, Dionisio Acosta, Juan M García-Gómez, Carlos Sáez

## Abstract

**Objectives:** To demonstrate how data-driven variability methods can be used to identify changes in disease recording in two English electronic health records databases between 2001-2015.

**Design:** Repeated cross-sectional analysis that applied data-driven temporal variability methods to assess month-by-month changes in routinely-collected medical data. A measure of difference between months was calculated based on joint distributions of age, gender, socio-economic status and recorded cardiovascular diseases. Distances between months were used to identify temporal trends in data recording.

**Setting:** 400 English primary care practices from the Clinical Practice Research Datalink (CPRD GOLD) and 451 hospital trusts from the Hospital Episode Statistics (HES).

**Main outcomes:** The proportion of patients (CPRD GOLD) and hospital admissions (HES) with a recorded cardiovascular disease (CPRD GOLD: coronary heart disease, heart failure, peripheral arterial disease, stroke; HES: International Classification of Disease ICD codes I20-I69/G45).

**Results:** Both databases showed gradual changes in cardiovascular disease recording between 2001 and 2008. The recorded prevalence of included cardiovascular diseases in CPRD GOLD increased by 47%-62%, which partially reversed after 2008. For hospital records in HES, there was a relative decrease in angina pectoris (−34.4%) and unspecified stroke (−42.3%) over the same time period, with a concomitant increase in chronic coronary heart disease (+14.3%). Multiple abrupt changes in the use of myocardial infarction codes in hospital were found in March/April 2010, 2012 and 2014, possibly linked to updates of clinical coding guidelines.

**Conclusions:** Identified temporal variability could be related to potentially non-medical causes such as updated coding guidelines. These artificial changes may introduce temporal correlation among diagnoses inferred from routine data, violating the assumptions of frequently used statistical methods. Temporal variability measures provided an objective and robust technique to identify, and subsequently account for, those changes in electronic health records studies without any prior knowledge of the data collection process.

## 1 Introduction

Routinely-collected electronic health records (EHR) are increasingly used for clinical research [1, 2]. They often pool data from different healthcare sites over multiple years, providing a readily-available and representative national sample of clinical practice. The validity of results from observational studies heavily depends on the quality of the data [3], and researchers have become increasingly aware of the importance of adequate data quality for obtaining reliable and reproducible findings [4, 5]. Systematic approaches to ascertain data quality in health data repositories have traditionally focused on the data quality dimensions of completeness, correctness, and concordance [6]. For example, validation studies of English EHR databases commonly focused on whether all relevant information on the patient is recorded (completeness), to which degree the recorded information reflects reality (correctness), and whether the recorded information agrees with information in a reference dataset (concordance) [7]. While answering these questions is of vital importance, they are not the only potential sources of bias.

Other factors that influence the reuse of data are less obvious and have often been neglected despite their potentially large impact on study results. Changes in clinical procedures over time, differences in processes between healthcare sites and the introduction of new guidelines can all cause unwarranted variations in the way data items are recorded, leading to artificial trends, irregularities and breaks in the data distributions. We have previously argued that these types of data variability over time and between participating healthcare sites pose a considerable threat to the validity and reproducibility of EHR studies [8]. Studies that ignore these variations are susceptible to obtain results of limited applicability. For example, financial incentives to improve diabetes care might exaggerate an increase in recorded and reported type II diabetes [9]. At worst, variability in how data is collected might even introduce spurious relationships, such as when the above mentioned improved coding of type II diabetes reduces the incidence of patients with wrongly classified type I diabetes [10].

In this study, we set out to demonstrate how data-driven methods can be used to identify irregularities in coding for clinical diagnoses over time using a recently developed, scalable approach that allows for easy comparison of similarities and differences in the distribution of demographic patient characteristics and cardiovascular diagnosis codes [8, 11]. Using this method, we show how changes in coding guidelines can and have affected cardiovascular disease recording in two major English EHR databases in primary care (Clinical Practice Research Datalink - CPRD GOLD [12]) and secondary care (Hospital Episode Statistics - HES [13]) and discuss potential causes of detected variations in coding over time.

## 2 Methods

### 2.1 Data Sources

#### Clinical Practice Research Datalink (CPRD GOLD)

CPRD GOLD is a database of retrospective health records obtained directly from the practice management software (Vision, InPractice Systems LTD) of 674 primary care practices across the UK [12]. Recorded information include patient’s demography, clinical symptoms, investigations, diagnoses, and tests entered by the clinician. All clinical information is coded using Read Codes, the clinical terminology used in UK primary care [14]. As of 2015, CPRD GOLD collected data from 674 practices, including data on 4.4 million actively contributing patients and 6.9 million historic patient records [12]. Data from CPRD GOLD used in this study was obtained as part of the Preserving Antibiotics through Safe Stewardship (PASS) project, a project investigating the association between comorbidity status and antibiotic prescribing [ref]. For consistency with the secondary care data, findings presented here represent all quality assurance measures performed for cardiovascular comorbidities only. Due to the scope of PASS, primary care data in this study was used exclusively from the subset of practices linked to hospital and census data. All data was obtained via the CALIBER research resource [15].

#### Hospital Episode Statistics (HES)

Hospital Episode Statistics (HES) Admitted Patient Care (APC) is a repository of hospital activity data collected as part of management, planning and reimbursement of NHS hospitals in England [13]. Information is organised in finished consultant episodes (i.e. the time spent under the uninterrupted care of a single consultant) and includes the patient’s demography, admission and discharge dates, hospital diagnoses, and performed procedures. Each episode has an assigned primary diagnosis, which denotes the main condition treated during that episode, and up to 19 secondary diagnoses that contain any comorbidities relevant to the episode. Diagnoses are coded using the International Classification of Diseases 10th revision (ICD-10) codes and surgical procedures are recorded using OPCS-4 codes. In the financial year 2014/15, a total of 18.7 million episodes from 451 NHS hospital trusts were captured in HES, which was equal to 34.3 episodes per 100 person-years [13]. Data from HES used in this study was provided by Public Health England and was pre-aggregated by month, age group, gender, socio-economic status and 3-character ICD-10 chapter.

### 2.2 Study design and population

We conducted a cross-sectional study of electronic health records from English primary care (CPRD GOLD) and secondary care (HES) between 2001 and 2015. The data from each database was divided into monthly cross-sectional slices (Figure 1 - A).

**Figure 1:**
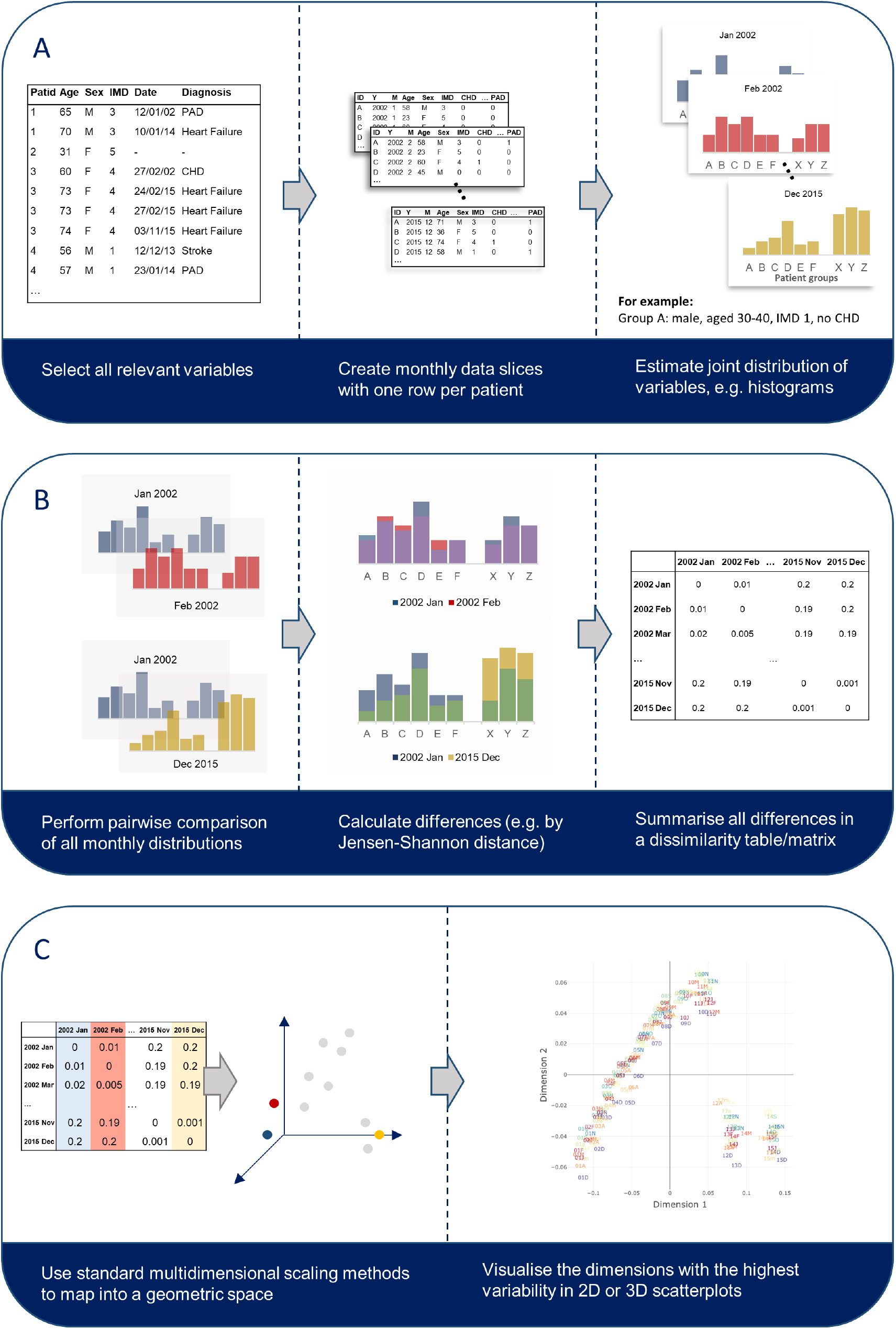
Step-by-step explanation to estimate and visualise the temporal variability of a dataset. Methods included in the R package EHRtemporalVariability were created to support researchers with all steps in this process.

For CPRD GOLD, patients were included in the batch for each month if they were between 20-110 years old at the beginning of the month and had been registered with their GP for at least one year. Patients and practices were further required to fulfill standard data quality checks as performed and reported by CPRD [12]. Data for each month included age, gender, socio-economic status (quintiles of the patient’s Index of Multiple Deprivation - IMD 2015 [16]), and presence of coronary heart disease (CHD), heart failure, peripheral arterial disease (PAD), and stroke. All cardiovascular conditions were separately ascertained at the first day of each month. Presence of cardiovascular disease was defined as the presence of a relevant diagnosis code at any time in the patient’s medical history in CPRD GOLD (see Supplementary Table 1). Included diagnosis codes were directly taken from the Quality and Outcomes Framework (QOF; version 36.0), a financial incentive scheme introduced in 2004 aimed at improving the management and recording of chronic disease in primary care. An absence of a diagnosis code in a patient’s medical history was interpreted as absence of the disease.

For HES, all admissions with a recorded diagnosis of ICD chapters I20-I69 (including CHD, heart failure, stroke, but not PAD; Supplementary Table 2) and G45 (transient ischaemic attacks) in patients aged 40 years or more were counted by month and stratified by age, gender and socioeconomic status (IMD 2015). We did not distinguish between primary and secondary diagnoses and included all codes recorded during an admission. A summary of the included data for each dataset can be found in Supplementary Tables 2-5.

### 2.3 Temporal variability metrics

Variation in coding across months was assessed in both datasets via temporal variability methods previously proposed and tested by two of the authors [8, 17]. These metrics are based on a comparison of the occurrence of a single trait (e.g. heart failure) or the joint occurrence of multiple traits (e.g. male and heart failure) within a patient population over time. The variability plots are calculated using a distribution or joint distribution, respectively, of all variables of interest for each month. In the simplest case where all variables are categorical, the joint distribution is a histogram of all possible value combinations (Figure 1 - A). Temporal variability quantifies the differences in those histograms/distributions across time intervals. The estimates of variability are based on the pairwise distance between each pair of monthly distributions (Figure 1 - B). For the purpose of this study we used the Jensen-Shannon distance (JSD), an information theoretic measure that estimates the degree of similarity between two probability distributions [11], where 0 means equal distributions, and 1 means no-overlap on the distributions. Notably, all pairwise distances are bound and independent of sample size. The dissimilarity matrix resulting from all pairwise comparisons was mapped into a Euclidean space using multidimensional scaling (Figure 1 - C; [18]). The resulting plot visualises the data’s evolution over time and allowed a graphical analysis of data recording trajectories, i.e. systematic patterns in the data evolution. A detailed description of the methods can be found in Sáez et al. (2015) [17] and Sáez et al. (2016) [8]. Functions to perform the temporal variability analysis were implemented using the R package EHRtemporalVariability (https://github.com/hms-dbmi/EHRtemporalVariability).

### 2.4 Statistical analysis

Empirical probability distributions were estimated for each month by calculating the joint histogram divided by the total number of observations in each month, i.e. the proportions of observations with each possible combination of variables. For CPRD GOLD, this was equivalent to the prevalence of cardiovascular diseases at the beginning of each calendar month. For HES, proportions represented the relative frequency of each included 3-character cardiovascular ICD-10 code (e.g. I21 Acute Myocardial Infarction). The denominator was the total number of cardiovascular codes recorded during a hospital admission. Proportions in both cases were stratified by age group, gender and socioeconomic status.

For each dataset, the temporal variability was calculated jointly for all covariates in a given month as described above. The estimated variability was plotted in a 3D scatter plot and visually inspected for data recording trajectories. Plots were searched for gradual trends, abrupt changes, seasonality, distinct subgroups, and outliers [19]. Where trends, breaks or discontinuities were observed, the same analysis was performed for each variable individually to investigate the reasons for the discovered deviation. Results

## 3 Results

The variables extracted from CPRD GOLD showed a gradual trend from one month to the next between 2001 and 2007 (Figure 2), mostly driven by changes in prevalence of cardiovascular diseases. The pattern suggested a continuous evolution of disease prevalence compatible with social factors (e.g. ageing) or incremental improvements in diagnostic coding or in clinical procedures. Smaller deviations from this overall trend could be seen at the end of 2002 and throughout 2005. Across the 8 years, the data distribution of cardiovascular disease prevalence changed with an average magnitude of about 1.5 * 10^−3^ JSD/month, which can be roughly viewed as a 0.15% difference between consecutive histograms of disease prevalence. This is comparable to a monthly increase in the prevalence of a single disease from 1% to 2.2% or from 50% to 55% (the magnitude of the estimated JSD depends on the base prevalence). Changes across this period were mainly attributable to an increase in the number of patients with heart failure (from 6.7/1000 patients at the start of 2001 to 10.8/1000 by the end of 2007; +62%), stroke (from 14.4/1000 to 23.4/1000; +62%), PAD (from 7.0/1000 to 10.3/1000; +47%), and to a lesser extent CHD (from 44.7/1000 to 48.2/1000; +7.8%).

**Figure 2:**
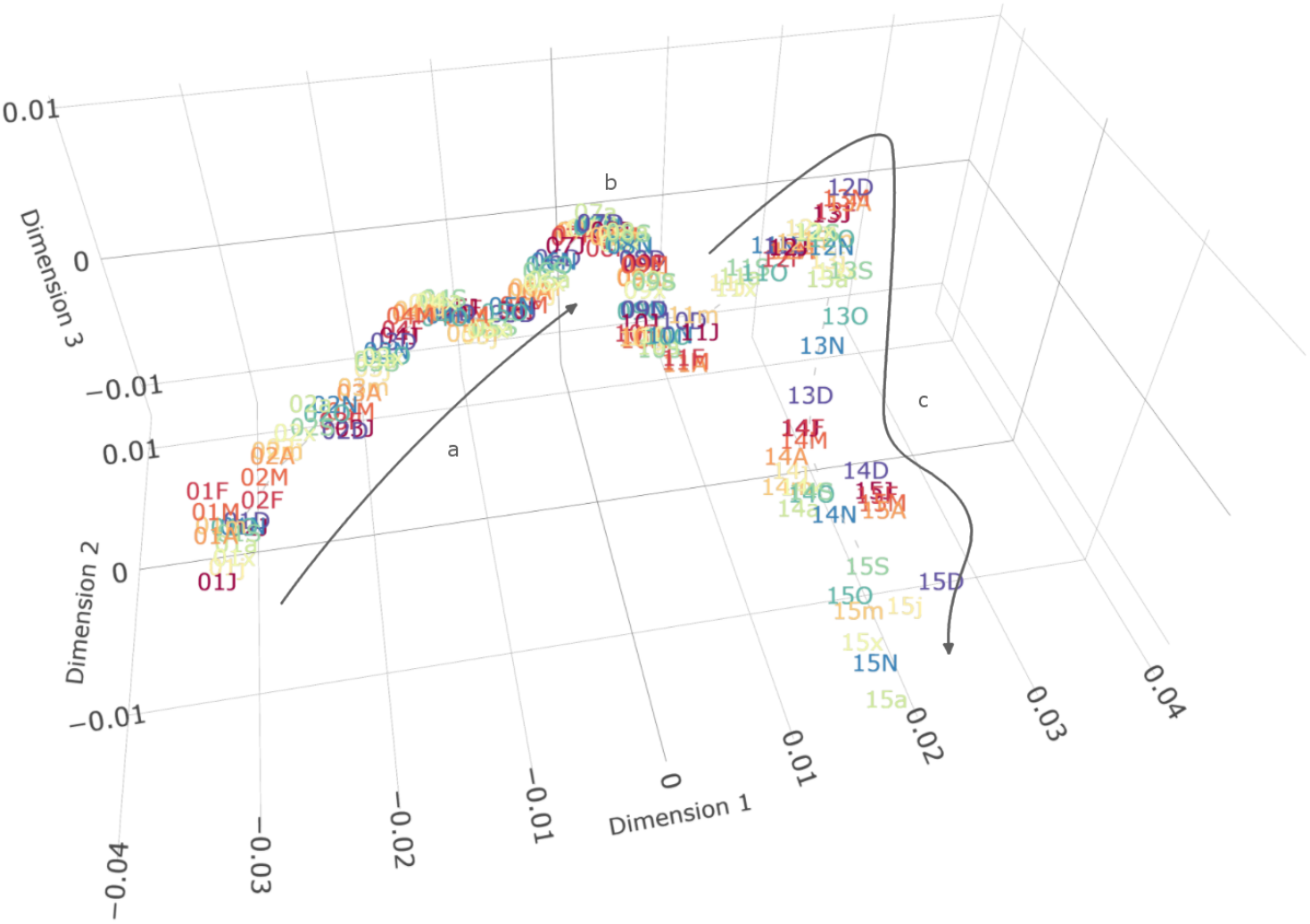
Information-Geometric Temporal (IGT) plot of demography (without age*) and cardiovascular disease prevalence in CPRD between 2001 and 2015. Each point represents joint prevalence in a single month (labelled with the last 2 digits of the year and month as given in the list of abbreviations) and distances represent the relative difference between them. Dimensions have no inherent meaning but represent the three ordered dimensions of highest variability as determined by multidimensional scaling. (A) Between 2001-2008 there was a gradual increase in disease prevalence, with two indentations corresponding to the years 2003 and 2005. (B) In 2008, the general trend reverses and prevalences decrease again, shown by a change in the direction of the graph. (C) The magnitude of variability increases after 2011, predominantly owing to changes in the socio-economic status due to a reduction in the number of practices contributing to the dataset. * The given graph excluded the age variable for clarity. Since CPRD GOLD includes only the year of birth, including age leads to artificial yearly jumps in July when every patient is considered one year older. The overall conclusion remains unaltered. A full graph including age can be found in the supplementary material.

From 2008 onwards the trend shifted direction, owing to a reduction and partial reversal in the prevalence of heart failure, PAD, and most notably CHD (from 49/1000 at the beginning of 2008 to 39/1000 at the end of 2015; 20% reduction). The estimated average magnitude of change increased to 2.4 * 10^−3^ JSD/month between 2008 and 2015. Starting in March 2011, the gradual pattern diverged from the relatively straight path seen before due to shifts in the socio-economic distribution of the patient population. This coincided with a substantial drop of contributing practices from more than 343 practices in January 2011 to 165 active practices in December 2015 (Supplementary Figure 1).

The distribution of cardiovascular diagnoses associated with HES admissions experienced a gradual change similar to that observed for CPRD GOLD until the end of 2008 (Figure 3). In this period, the use of codes generally stayed comparable and shifted only over the course of multiple years. Notable changes were seen for I20 Angina pectoris (- 34.4%), I64 Unspecified stroke (−42.3%) and I25 Chronic CHD (+14.3%). After a transition period in 2009, distributions stopped to evolve gradually and started to cluster tightly by NHS financial year (April to March - Figure 3). There was a major shift in cardiovascular admission coding every two years (financial years 2010/11, 2012/13 and 2014/15). The abrupt changes were primarily due to differences in the ICD-10 codes used, while age, gender and socioeconomic status remained largely stable.

**Figure 3:**
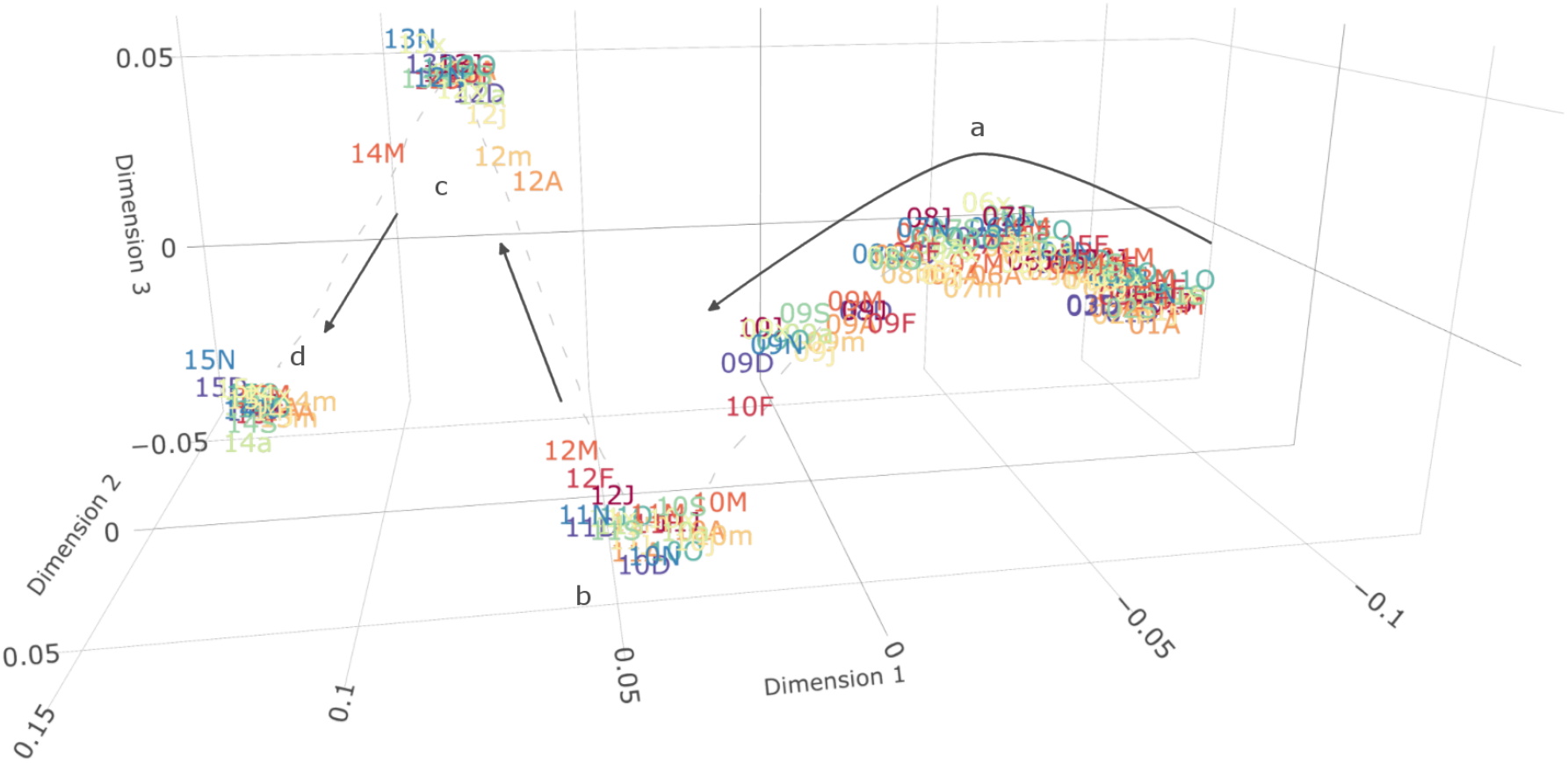
Information-Geometric Temporal (IGT) plot of demography and cardiovascular disease coding in HES between 2001 and 2015. Each point represents joint prevalence in a single month (labelled with the last 2 digits of the year and month as given in the list of abbreviations) and distances represent the relative difference between them. Dimensions have no inherent meaning but represent the three dimensions of highest variability (in order) as determined by multidimensional scaling. (A) From 2001-2009 there was a gradual change in which cardiovascular codes were associated with hospital admission. The data distributions starts to diverge from the previous trend in March 2009. (B) In March 2010, the distribution of cardiovascular codes abruptly changes. (C and D) Similar and even stronger changes in cardiovascular disease coding occurred again in April 2012 and April 2014. The distributions within these 2-year batches remained stable.

In particular, chapters I20-I25 experienced noticeable temporal breaks (Figure 4). While I21 Acute myocardial infarction declined in relative frequency starting in 2006, it increased from 8.0% of included codes in March 2012 to 10.9% in April 2012 (+36%) and remained stable thereafter. Simultaneously, the related code I22 Subsequent myocardial infarction (including reinfarction and recurrent infarction) dropped from 1.4% in March to 0.5% in April (−64%) and finally to 0.1% after September 2012. I20 Angina pectoris decreased from 18.8% in January 2001 to 10.2% in March 2014 (−45.7%), after which it further declined by 2% points to 8.4% in April 2014 (−17.6%).

**Figure 4:**
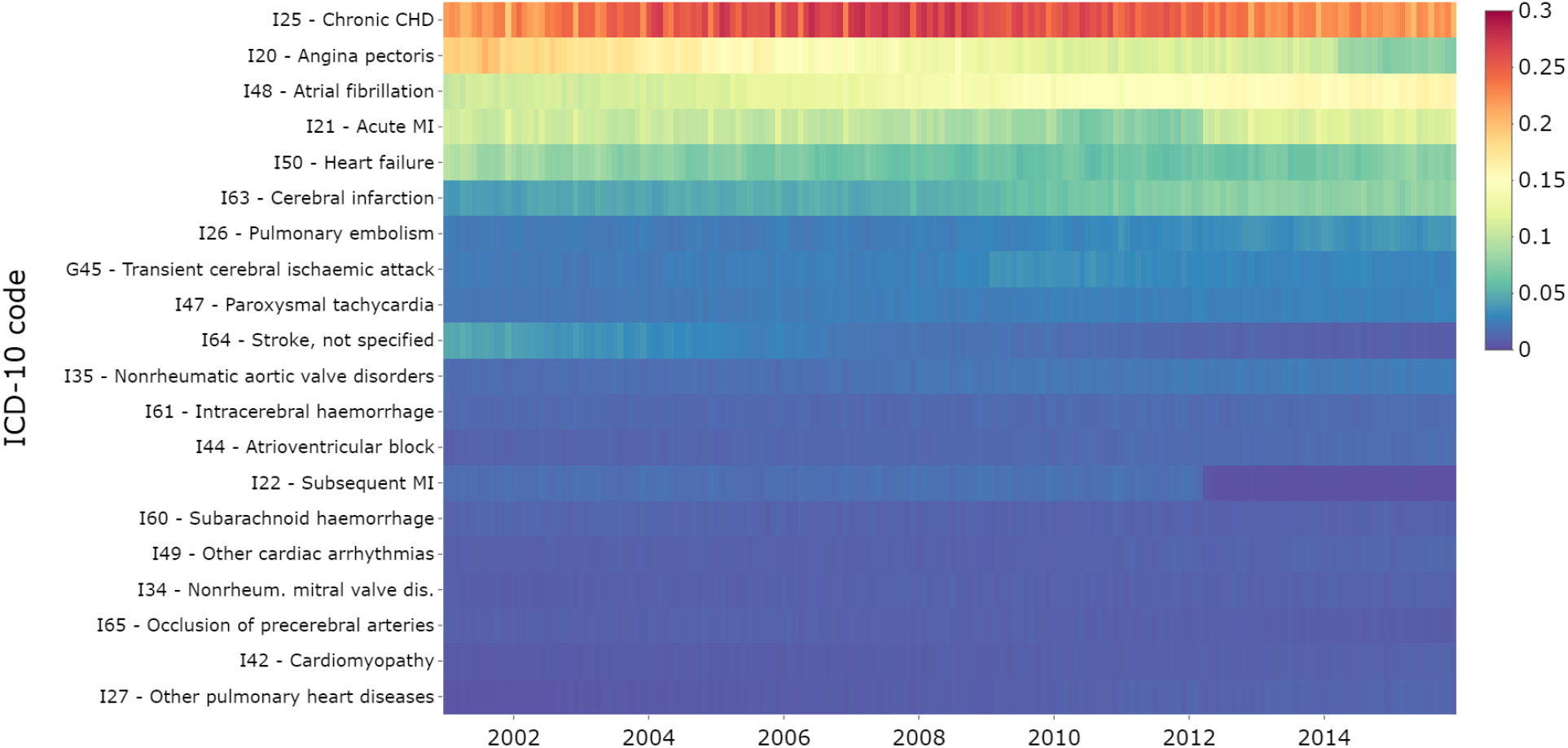
Data Temporal Map (DTM) of ICD-10 coding linked to hospital admissions in HES between 2001 and 2015. Each row represents a single ICD-10 code (3 characters) and the colour shows the proportion of admissions with that code in each month. Gradual changes in code frequency can notably be seen for I20 - Angina Pectoris, I21 - Acute Myocardial Infarction, I63 - Cerebral Infarction, and I64 - Stroke, not specified. Abrupt changes appear in the coding of G45 - Transient Cerebral ischaemic attack (2009), I21 - Acute Myocardial Infarction (2010 and 2012), and I20 - Angina Pectoris (2014).

## 4 Discussion

We discovered both gradual and abrupt changes in the distribution of cardiovascular patient populations in two large English EHR databases between 2001 and 2015 using recently developed data quality measures (Table 1). The observed differences in cardiovascular disease coding might bias clinical phenotypes when applied over the entire study period, introducing correlation within time periods that violate the assumptions underlying common statistical methods (e.g. regression analysis). Temporal variability measures provided an objective and robust way to identify those changes without any prior knowledge of the data.

**Table 1:**
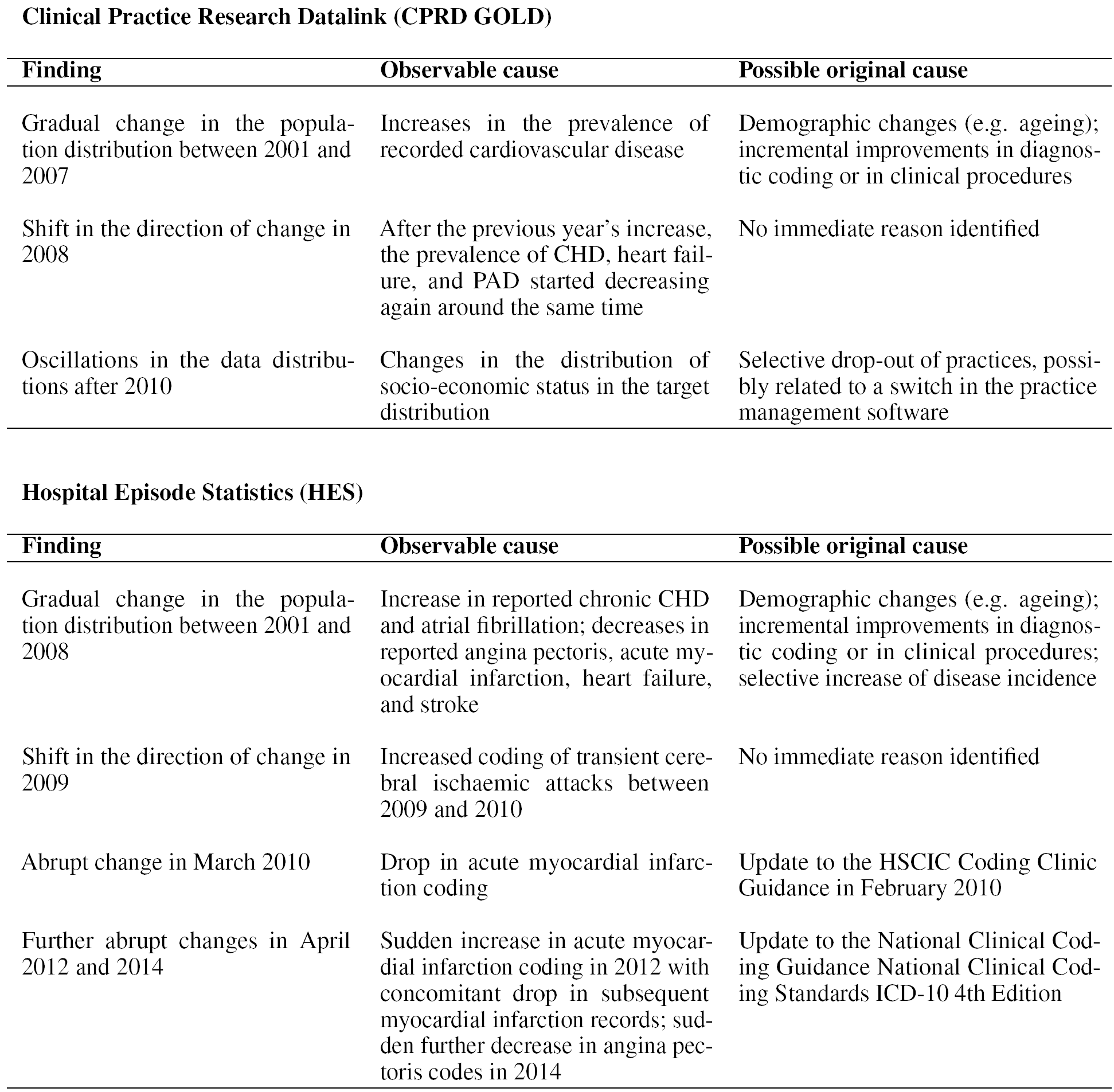
Variability in the Clinical Practice Research Datalink (CPRD) and Hospital Episode Statistics (HES) and their potential causes

**Clinical Practice Research Datalink (CPRD GOLD)**
| **Finding** | **Observable cause** | **Possible original cause** |
| --- | --- | --- |
| Gradual change in the population distribution between 2001 and 2007 | Increases in the prevalence of recorded cardiovascular disease | Demographic changes (e.g. ageing); incremental improvements in diagnostic coding or in clinical procedures |
| Shift in the direction of change in 2008 | After the previous year's increase, the prevalence of CHD, heart failure, and PAD started decreasing again around the same time | No immediate reason identified |
| Oscillations in the data distributions after 2010 | Changes in the distribution of socio-economic status in the target distribution | Selective drop-out of practices, possibly related to a switch in the practice management software |

**Hospital Episode Statistics (HES)**
| **Finding** | **Observable cause** | **Possible original cause** |
| --- | --- | --- |
| Gradual change in the population distribution between 2001 and 2008 | Increase in reported chronic CHD and atrial fibrillation; decreases in reported angina pectoris, acute myocardial infarction, heart failure, and stroke | Demographic changes (e.g. ageing); incremental improvements in diagnostic coding or in clinical procedures; selective increase of disease incidence |
| Shift in the direction of change in 2009 | Increased coding of transient cerebral ischaemic attacks between 2009 and 2010 | No immediate reason identified |
| Abrupt change in March 2010 | Drop in acute myocardial infarction coding | Update to the HSCIC Coding Clinic Guidance in February 2010 |
| Further abrupt changes in April 2012 and 2014 | Sudden increase in acute myocardial infarction coding in 2012 with concomitant drop in subsequent myocardial infarction records; sudden further decrease in angina pectoris codes in 2014 | Update to the National Clinical Coding Guidance National Clinical Coding Standards ICD-10 4th Edition |

Many studies have used CPRD GOLD to look at the incidence [20, 21, 22, 23], prevalence [20, 21, 23, 24] and outcomes of cardiovascular disease [25, 26]. However, none of these studies mentioned changes in the coding of diseases, and only one of these studies reported findings per year [23]. Changes in coding over time tend to be underreported in research papers, their identification limited to dedicated validation studies, which depending on the disease investigated may or may not exist. To the best of our knowledge, no patterns in the recording of included cardiovascular diseases have been recorded for CPRD yet. Previous validation studies in other chronic diseases did report changes in coding over time. Among others, improved coding has been reported for chronic obstructive pulmonary disease [27], diabetes [10], body mass index [28], and smoking status [29]. Improvements in primary care coding were primarily linked to the introduction of QOF in 2004, a payment scheme which incentivised better coding and management of chronic diseases. In line with these studies, we found alterations of the overall trend leading up to QOF (December 2002 - December 2003) and following its introduction (October 2004 - September 2005). However, these effects were small and the estimated prevalence had been already rising before 2003 and kept increasing after 2005. A study analysing the coding of diabetes further found a sharp increase of coded type 2 diabetes in 2004 that slowly started to decrease again after 2008 [9], around the same time that we observed a reversal of cardiovascular prevalence. It is unclear what prompted these changes, and whether they might be related. Our method further detected notable changes in patient population after 2011, mainly due to changes in the distribution of socio-economic status. This was likely related to a considerable reduction in the number of participating practices from around 350 to 165 at the end of the study period, potentially due to practices switching to new practice management software incompatible with CPRD GOLD [30].

Data on cardiovascular admissions from HES have been used alone or linked with primary care data from CPRD GOLD [24]. Again, to the best of our knowledge, no changes in cardiovascular disease coding in HES have been published previously. A validation study comparing coronary heart disease in HES (ICD-10 codes I20-I25) to data from the prospective UK Whitehall II cohort study [31] found a generally good agreement between the two data sources [32], suggesting reasonable recording quality in HES. An earlier study comparing data from HES and the Million Women Study [33] came to a similar conclusion [34]. However, the studies only included data up to 2013 and 2005 respectively, and did not compare data from different time periods. Other studies based on local audits or comparison to specific disease registries [35] have reported a notable underreporting of myocardial infarction cases in HES [21, 36]. Any increases or decreases in the observed number of cardiovascular disease could thus be due to improvements or deteriorations in coding. The gradual change in distributions observed until 2008 agreed with the trends observed in CPRD GOLD and could either relate to more/less focused coding for certain diseases or a slow shift in the characteristics of the underlying patient populations. The first major change occurred in March 2010, following a new version of NHS Digital / HSCIC Coding Clinic Guidance in February [37]. This guideline included provisions for stricter coding of I21 Acute Myocardial Infarction, requiring a different coding of myocardial infarction in subsequent trusts to avoid overcounting. The additional changes in 2012 and 2014 both happened in April, coinciding with the financial year of the National Health Service and the publishing of updates to the National Clinical Coding standards [38], making it likely that they too are the result of changes in coding practice. As these changes mostly occurred within the group I20-I25, they might not affect studies that use all of these codes, but may lead to problems if authors include only a single code from this group (e.g. I21). Related preliminary results on hospital admissions for all ICD-10 codes (not only chapter I) using a traditional interrupted time series analysis showed further, similar changes in non-cardiovascular chapters; these results will be disseminated in a further study evaluating life-style related diseases.

It is challenging to disentangle changes solely due to how diseases are recorded from other, genuine shifts in the patient population. While abrupt changes like the one observed here for myocardial infarction strongly suggest an exogenous cause such as new clinical coding guidelines, continuous, gradual changes over a long time period can be more difficult to classify. However, we believe that is important that researchers are aware of potential variations irrespective of the cause. Even in cases where changes are attributable to demographic shifts, accounting for them in the statistical analysis might still be warranted. The impact of observed changes, genuine as well as artificial, always depends on the research question under investigation [39]. For example, an increase in the estimated population prevalence of heart failure from 0.7% to 1.0% might not impact findings when accounting for heart failure as a covariate in a larger cohort, but might significantly alter the patient characteristics in a smaller, heart failure-only cohort. Insights gained from temporal variability analysis can be used to investigate and account for changes in patient cohorts across years.

Although some of the findings presented in this study could be detected with conventional methods such as traditional time series analysis of incidence rates, these methods usually require a formal definition of the time point at which changes happen. They further do not handle multivariate, multitype and multimodal data well [17] and require a separate analysis for each variable. This is particularly problematic when analysing changes in multinomial variables such as ICD-10 codes. More traditional methods might further struggle with large sample sizes, whereas the structure of the variability metrics allows for a flexible modelling and subsequent hypothesis testing via statistical process control. Temporal variability metrics together with the tools provided in our EHRtemporalVariability R package facilitate the calculus from raw data tables directly to visualization. Results can be shared on the Shiny user interface (http://ehrtemporalvariability.upv.es), aiding transparency and communication.

### 4.1 Limitations

Results in this study are limited by the fact that the conditions chosen for inclusion represented a convenience sample based on the overlap between the two projects for which the data was originally obtained. The results shown here therefore do not constitute a systematic, in-depth validation study of cardiovascular disease recording. Indeed, the aim of this study was not to comprehensively investigate the data quality of cardiovascular coding in CPRD GOLD and HES but to show how systematic, data-driven methods for studying temporal variability can help to identify potential coding inconsistencies over time early on in a project and allow researchers to a priori adjust the analysis accordingly. We believe that routine checks of the temporal variability of study data will aid the validity and reproducibility of medical and epidemiological studies. Reporting coding variability in supplementary material can help readers judge the reliability of codelists and strengthen the conclusions. The analysis presented here was also limited to a manual inspection of the plots, as would be appropriate for interactive data quality analysis at the start of a project. The framework can easily be extended to allow for a more formal statistical process control (see Sáez et al. (2015 and 2018) [17, 19] for guidance).

Despite the promising results, we must note that variability metrics are solely based on recorded data and will not detect the same data quality issues and trends identified by other, dedicated validation studies based on manual code review or GP questionnaires [7]. They are not meant to replace in-depth validation of data sources but rather complement them. Extensive validation studies are costly and are dependent on the exact codelists used during validation. Our data-driven approach might be well suited as a first step to signal the need for an extensive validation. With regards to the findings in this study, a reasonable first step in assessing their impact for a specific research study might be to perform analysis stratified by NHS financial year or observed stable periods. How results should be reported and whether the data is fit for purpose then depends on the results of this sensitivity analysis and the exact research questions investigated.

### 4.2 Conclusion

We identified previously unreported variability in the frequency of cardiovascular codes in CPRD GOLD and HES between 2001 and 2015 using temporal variability measures that require minimal prior specification. In doing so, we have demonstrated the utility of application of data-driven approaches to data quality on two of the most important data resources for clinical research in the UK. We demonstrated that the methods can be implemented in an unsupervised, scalable manner, providing non-parametric visualisations of data recording trajectories to measure their variability. The results from this variability analysis enable researchers to adjust their analysis and ensure reproducible results.

## Data Availability

CPRD data cannot be directly shared by the researchers but are available directly from CPRD subject to standard conditions. [HES data sharing disclaimer]. All statistical code is available from https://github.com/prockenschaub/CPRD_variability.

https://github.com/prockenschaub/CPRD_variability

## Acknowledgements

We thank Dr Anoop Shah for their invaluable comments on the manuscript draft. This study is based in part on data from the Clinical Practice Research Datalink obtained under license from the UK Medicines and Healthcare products Regulatory Agency. The data are provided by patients and collected by the NHS as part of their care and support. Hospital Episode Statistics Copyright (2019) is reused with the permission of The Health & Social Care Information Centre. All rights reserved. All primary care data was obtained through CALIBER, which is a research resource consisting of linked electronic health records phenotypes, methods and tools, specialized infrastructure, and training and support led from the UCL Institute of Health Informatics. The conclusions and views expressed are those of the author(s) and not necessarily those of the National Health Service, Public Health England, or the Department of Health.

## Ethical approval

The primary care analysis reported in this study was approved by the Medicines and Healthcare products Regulatory Agency’s independent scientific advisory committee (protocol 17 048). For analysis of secondary care data, the authors had only access to the aggregated data which was provided by Public Health England. No separate ethical approval was required.

## Data sharing

CPRD data cannot be directly shared by the researchers but are available directly from CPRD subject to standard conditions. Similarly, HES data too cannot be shared by the researchers but is available from NHS Digital subject to standard conditions. All statistical code is available from https://github.com/prockenschaub/CPRD_variability.

## Funding statement

This work was supported by the Economic and Social Research Council (ES/P008321/1). JMGG and CS contributions to this work were partially supported by the MTS4up Spanish project (National Plan for Scientific and Technical Research and Innovation 2013-2016, No. DPI2016-80054-R), the CrowdHealth H2020-SC1-2016-CNECT project (No. 727560) (JMGG) and the Inadvance H2020-SC1-BHC-2018-2020 project (No. 825750).

## Competing interests

None declared

## Notes

### Competing Interest Statement

The authors have declared no competing interest.

### Author Declarations

All relevant ethical guidelines have been followed and any necessary IRB and/or ethics committee approvals have been obtained.

Any clinical trials involved have been registered with an ICMJE-approved registry such as ClinicalTrials.gov and the trial ID is included in the manuscript.

## References

[1] George Hripcsak and David J Albers. Next-generation phenotyping of electronic health records. J. Am. Med. Inform. Assoc., 20(1):117–121, January 2013.

[2] Paul R Burton, Madeleine J Murtagh, Andy Boyd, James B Williams, Edward S Dove, Susan E Wallace, Anne-Marie Tassé, Julian Little, Rex L Chisholm, Amadou Gaye, Kristian Hveem, Anthony J Brookes, Pat Goodwin, Jon Fistein, Martin Bobrow, and Bartha M Knoppers. Data safe havens in health research and healthcare. Bioinformatics, 31(20):3241–3248, October 2015.

[3] Ricardo Cruz-Correia, Pedro Rodrigues, Alberto Freitas, Filipa Almeida, Rong Chen, and Altamiro Costa-Pereira. Data quality and integration issues in electronic health records, 2009.

[4] Barbara L Massoudi, Kenneth W Goodman, Ivan J Gotham, John H Holmes, Lisa Lang, Kathleen Miner, David D Potenziani, Janise Richards, Anne M Turner, and Paul C Fu. An informatics agenda for public health: summarized recommendations from the 2011 AMIA PHI conference. J. Am. Med. Inform. Assoc., 19(5):688–695, September 2012.

[5] D R Schlegel and G Ficheur. Secondary use of patient data: Review of the literature published in 2016. Yearb. Med. Inform., 26(1):68–71, August 2017.

[6] Nicole Gray Weiskopf and Chunhua Weng. Methods and dimensions of electronic health record data quality assessment: enabling reuse for clinical research. J. Am. Med. Inform. Assoc., 20(1):144–151, January 2013.

[7] Emily Herrett, Sara L Thomas, W Marieke Schoonen, Liam Smeeth, and Andrew J Hall. Validation and validity of diagnoses in the general practice research database: a systematic review. Br. J. Clin. Pharmacol., 69(1):4–14, January 2010.

[8] Carlos Sáez, Oscar Zurriaga, Jordi Pérez-Panadés, Inma Melchor, Montserrat Robles, and Juan M García-Gómez. Applying probabilistic temporal and multisite data quality control methods to a public health mortality registry in spain: a systematic approach to quality control of repositories. J. Am. Med. Inform. Assoc., 23(6):1085–1095, 2016.

[9] A Rosemary Tate, Sheena Dungey, Simon Glew, Natalia Beloff, Rachael Williams, and Tim Williams. Quality of recording of diabetes in the UK: how does the GP’s method of coding clinical data affect incidence estimates? cross-sectional study using the CPRD database. BMJ Open, 7(1):e012905. January 2017.

[10] Melanie Calvert, Aparna Shankar, Richard J McManus, Helen Lester, and Nick Freemantle. Effect of the quality and outcomes framework on diabetes care in the united kingdom: retrospective cohort study. BMJ, 338:b1870. May 2009.

[11] Carlos Sáez, Montserrat Robles, and Juan M García-Gómez. Stability metrics for multi-source biomedical data based on simplicial projections from probability distribution distances. Stat. Methods Med. Res., 26(1):312–336, February 2017.

[12] Emily Herrett, Arlene M Gallagher, Krishnan Bhaskaran, Harriet Forbes, Rohini Mathur, Tjeerd van Staa, and Liam Smeeth. Data resource profile: Clinical practice research datalink (CPRD). Int. J. Epidemiol., 44(3):827–836, June 2015.

[13] Annie Herbert, Linda Wijlaars, Ania Zylbersztejn, David Cromwell, and Pia Hardelid. Data resource profile: Hospital episode statistics admitted patient care (HES APC). Int. J. Epidemiol., 46(4):1093–1093i, August 2017.

[14] J Chisholm. The read clinical classification. BMJ, 300(6732):1092, April 1990.

[15] Spiros Denaxas, Arturo Gonzalez-Izquierdo, Kenan Direk, Natalie K Fitzpatrick, Ghazaleh Fatemifar, Amitava Banerjee, Richard J B Dobson, Laurence J Howe, Valerie Kuan, R Tom Lumbers, Laura Pasea, Riyaz S Patel, Anoop D Shah, Aroon D Hingorani, Cathie Sudlow, and Harry Hemingway. UK phenomics platform for developing and validating electronic health record phenotypes: CALIBER. J. Am. Med. Inform. Assoc., July 2019.

[16] Department for Communities and Local Government. The english index of multiple deprivation (IMD) 2015 - guidance. Technical report.

[17] Carlos Sáez, Pedro Pereira Rodrigues, João Gama, Montserrat Robles, and Juan M García-Gómez. Probabilistic change detection and visualization methods for the assessment of temporal stability in biomedical data quality. Data Min. Knowl. Discov., 29(4):950–975, July 2015.

[18] I Borg and P Groenen. Modern multidimensional scaling: Theory and applications. J Educational Measurement, 40(3):277–280, September 2003.

[19] Carlos Sáez and Juan M García-Gómez. Kinematics of big biomedical data to characterize temporal variability and seasonality of data repositories: Functional data analysis of data temporal evolution over non-parametric statistical manifolds. Int. J. Med. Inform., 119:109–124, November 2018.

[20] Nathalie Conrad, Andrew Judge, Jenny Tran, Hamid Mohseni, Deborah Hedgecott, Abel Perez Crespillo, Moira Allison, Harry Hemingway, John G Cleland, John J V McMurray, and Kazem Rahimi. Temporal trends and patterns in heart failure incidence: a population-based study of 4 million individuals. Lancet, 391(10120):572–580, February 2018.

[21] Emily Herrett, Anoop Dinesh Shah, Rachael Boggon, Spiros Denaxas, Liam Smeeth, Tjeerd van Staa, Adam Timmis, and Harry Hemingway. Completeness and diagnostic validity of recording acute myocardial infarction events in primary care, hospital care, disease registry, and national mortality records: cohort study. BMJ, 346:f2350. May 2013.

[22] Mar Pujades-Rodriguez, Adam Timmis, Dimitris Stogiannis, Eleni Rapsomaniki, Spiros Denaxas, Anoop Shah, Gene Feder, Mika Kivimaki, and Harry Hemingway. Socioeconomic deprivation and the incidence of 12 cardiovascular diseases in 1.9 million women and men: implications for risk prediction and prevention. PLoS One, 9(8):e104671. August 2014.

[23] Sally Lee, Anna C E Shafe, and Martin R Cowie. UK stroke incidence, mortality and cardiovascular risk management 1999-2008: time-trend analysis from the general practice research database. BMJ Open, 1(2):e000269. January 2011.

[24] Prachi Bhatnagar, Kremlin Wickramasinghe, Julianne Williams, Mike Rayner, and Nick Townsend. The epidemiology of cardiovascular disease in the UK 2014. Heart, 101(15):1182–1189, August 2015.

[25] Clare J Taylor, José M Ordóñez-Mena, Andrea K Roalfe, Sarah Lay-Flurrie, Nicholas R Jones, Tom Marshall, and F D Richard Hobbs. Trends in survival after a diagnosis of heart failure in the united kingdom 2000-2017: population based cohort study. BMJ, 364:223, February 2019.

[26] Johannes M I H Gho, Amand F Schmidt, Laura Pasea, Stefan Koudstaal, Mar Pujades-Rodriguez, Spiros Denaxas, Anoop D Shah, Riyaz S Patel, Chris P Gale, Arno W Hoes, John G Cleland, Harry Hemingway, and Folkert W Asselbergs. An electronic health records cohort study on heart failure following myocardial infarction in england: incidence and predictors. BMJ Open, 8(3):e018331. March 2018.

[27] Jennifer K Quint, Hana Müllerova, Rachael L DiSantostefano, Harriet Forbes, Susan Eaton, John R Hurst, Kourtney Davis, and Liam Smeeth. Validation of chronic obstructive pulmonary disease recording in the clinical practice research datalink (CPRD-GOLD). BMJ Open, 4(7):e005540. July 2014.

[28] Krishnan Bhaskaran, Harriet J Forbes, Ian Douglas, David A Leon, and Liam Smeeth. Representativeness and optimal use of body mass index (BMI) in the UK clinical practice research datalink (CPRD). BMJ Open, 3(9):e003389. September 2013.

[29] Helen P Booth, A Toby Prevost, and Martin C Gulliford. Validity of smoking prevalence estimates from primary care electronic health records compared with national population survey data for england, 2007 to 2011. Pharmacoepidemiol. Drug Saf., 22(12):1357–1361, December 2013.

[30] Helen Booth, Daniel Dedman, and Achim Wolf. CPRD aurum frequently asked questions (FAQs). Technical report, CPRD, April 2019.

[31] M G Marmot, G D Smith, S Stansfeld, C Patel, F North, J Head, I White, E Brunner, and A Feeney. Health inequalities among british civil servants: the whitehall II study. Lancet, 337(8754):1387–1393, June 1991.

[32] Mika Kivimäki, G David Batty, Archana Singh-Manoux, Annie Britton, Eric J Brunner, and Martin J Shipley. Validity of cardiovascular disease event ascertainment using linkage to UK hospital records. Epidemiology, 28(5):735–739, September 2017.

[33] P G Crosignani. Breast cancer and hormone-replacement therapy in the million women study. Maturitas, 46(2):91–92, October 2003.

[34] F Lucy Wright, Jane Green, Dexter Canoy, Benjamin J Cairns, Angela Balkwill, Valerie Beral, and Million Women Study Collaborators. Vascular disease in women: comparison of diagnoses in hospital episode statistics and general practice records in england. BMC Med. Res. Methodol., 12:161, October 2012.

[35] Emily Herrett, Liam Smeeth, Lynne Walker, Clive Weston, and MINAP Academic Group. The myocardial ischaemia national audit project (MINAP). Heart, 96(16):1264–1267, August 2010.

[36] L E Silver, C Heneghan, Z Mehta, A P Banning, and P M Rothwell. Substantial underestimation of incidence of acute myocardial infarction by hospital discharge diagnostic coding data: a prospective population-based study. Heart, 95(Suppl 1):2–2, June 2009.

[37] Health and Social Care Information Centre. Coding clinic guidance. Technical report, February 2010.

[38] Health & Social Care Information Centre. National clinical coding standards - ICD-10 4th edition. Technical report, February 2013.

[39] Richard Y Wang and Diane M Strong. Beyond accuracy: What data quality means to data consumers. Journal of Management Information Systems, 12(4):5–33, March 1996.

